# Reduced Neutralization of SARS-CoV-2 Omicron Variant by Vaccine Sera and Monoclonal Antibodies

**DOI:** 10.1101/2021.12.07.21267432

**Authors:** Alexander Wilhelm, Marek Widera, Katharina Grikscheit, Tuna Toptan, Barbara Schenk, Christiane Pallas, Melinda Metzler, Niko Kohmer, Sebastian Hoehl, Fabian A. Helfritz, Timo Wolf, Udo Goetsch, Sandra Ciesek

## Abstract

Due to numerous mutations in the spike protein, the SARS-CoV-2 variant of concern Omicron (B.1.1.529) raises serious concerns since it may significantly limit the antibody-mediated neutralization and increase the risk of reinfections. While a rapid increase in the number of cases is being reported worldwide, until now there has been uncertainty about the efficacy of vaccinations and monoclonal antibodies. Our *in vitro* findings using authentic SARS-CoV-2 variants indicate that in contrast to the currently circulating Delta variant, the neutralization efficacy of vaccine-elicited sera against Omicron was severely reduced highlighting T-cell mediated immunity as essential barrier to prevent severe COVID-19. Since SARS-CoV-2 Omicron was resistant to casirivimab and imdevimab, genotyping of SARS-CoV-2 may be needed before initiating mAb treatment. Variant-specific vaccines and mAb agents may be required to treat COVID-19 due to Omicron and other emerging variants of concern.

---

The SARS-CoV-2 variant Omicron was first identified in South Africa on November 9, 2021. Due to numerous mutations in the spike protein (S), which is the antigenic target of vaccine-elicited antibodies, Omicron raises serious concerns about significantly reduced vaccine efficacy and increased risk of reinfection^1^. Compared to the parental variant (B.1), Omicron S has 30 non-synonymous substitutions, three small deletions and an insertion (**Supplementary Figure 1, Supplementary Tables 1-3**). Fifteen of these mutations are in the receptor-binding domain (RBD), a major target of neutralizing antibodies (NAbs)^2^. Several of the S mutations observed in Omicron were reported in preceding variants of concern (VoCs) like Alpha, Beta, Gamma, Delta as well as variants of interest such as Kappa, Zeta, Lambda, and Mu (**Supplementary Table 3**) that were associated with higher transmissibility and immune escape. So far, Beta and Mu exhibited the most severe immune evading capacities^3,4^. Due to the high accumulation of these mutations in Omicron S, synergistic effects are expected and it is unclear whether prior immunity protects against re-infections.

To evaluate the protective capacity, antibody-mediated neutralization efficacy against authentic SARS-CoV-2 Omicron was determined *in vitro* using an isolate obtained from a double mRNA-1273-vaccinated travel returnee from Zimbabwe and compared to Delta. Neutralization performed with sera from double or triple BNT162b2-vaccinated individuals (6, 0.5 or 3 months after last vaccination/booster) revealed an 11.4-, 37.0- and 24.5-fold reduction, respectively (**Figure 1A**). Sera from double mRNA-1273-vaccinated and additionally BNT162b2-vaccinated individuals (sampled 6 or 0.5 months after last vaccination/booster) showed a 20- and 22.7-fold reduction in the neutralization capacity (**Figure 1B**). Poor neutralization against Delta and no efficacy against Omicron were observed using sera from heterologous ChAdOx1/BNT162b2-vaccinated individuals (**Figure 1C**). Additionally, the group receiving a third BNT162b2-vaccination showed a significant increase of NAb titers but a 27.1-fold reduction in neutralization against Omicron (**Figure 1C**). Neutralization of Omicron was 32.8-fold reduced using sera from double BNT162b2-vaccinated and previously SARS-CoV-2-infected individuals (**Figure 1A**).

**Figure 1.**
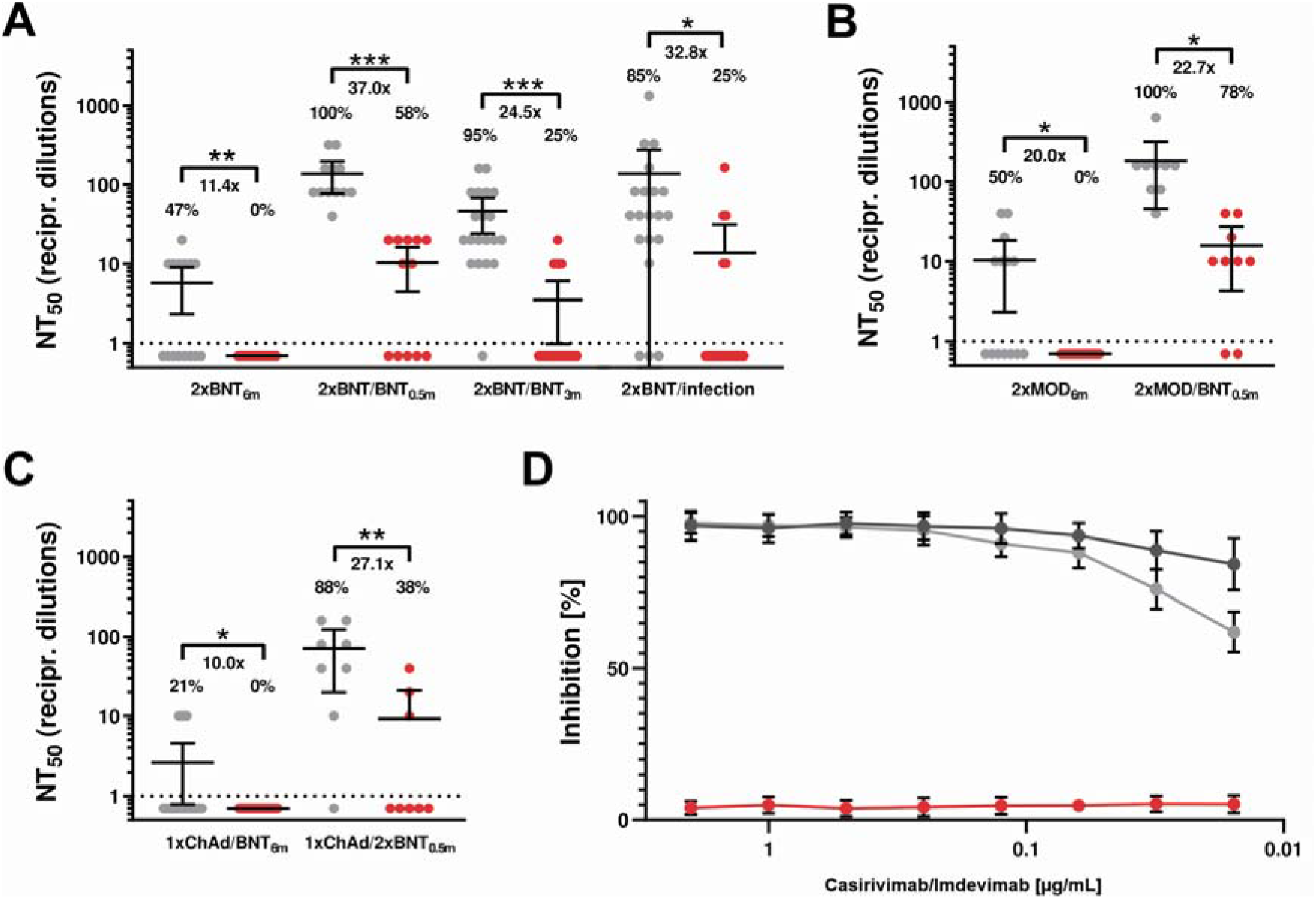
Antibody-mediated neutralization efficacy against authentic SARS-CoV-2 variants Delta and Omicron. Values represent reciprocal dilutions of SARS-CoV-2 variants Delta (grey) and Omicron (red) micro-neutralization titers resulting in 50% virus neutralization (NT_50_). **A)** Neutralization assays were performed using serum samples obtained from vaccinated individuals receiving the indicated vaccine schemes (sampling time after last vaccination/booster indicated in subscript): double BNT162b2-vaccinated (2xBNT_6m_), sera from triple BNT162b2-vaccinated individuals (2xBNT/BNT_0.5m_ and 2xBNT/BNT_3m_) and double BNT162b2-vaccinated and SARS-CoV-2 infected individuals (2xBNT/infection). **B)** Neutralization assays with sera from double mRNA-1273-vaccinated (2xMOD_6m_) and additionally BNT162b2-boosted (2xMOD/BNT_0.5m_). **C)** Neutralization titers for sera from heterologous ChAdOx1 and BNT162b2-vaccinated (1xChAd/1xBNT_6m_) and BNT162b2 boosted (1xChAd/2xBNT_0.5m_) individuals. The x-fold reduction was determined using the difference between NT_50_ values for Delta and Omicron. Only Delta neutralizing samples were considered and negative titers were handled as 1 for fold-change calculation. The percentages indicate the relative number of sera that yielded a measurable titer. Information regarding the sera donors (sex, age, antibody titers test and time after vaccination) are summarized in **Supplementary Table 4. D)** Neutralization efficacy of monoclonal antibodies imdevimab and casirivimab against SARS-CoV-2 Omicron (red), parental strain B (dark grey), and Delta (grey). The indicated concentrations of mAbs casirivimab and imdevimab were applied in a 1:1 ratio. Mean values of two technical replicates per sample are depicted with 95% confidence intervals and SD. All experiments were verified using a second SARS-CoV-2 Omicron strain (EPI_ISL_6959868; **Supplementary Figure 1**). Statistical significance compared to Delta was calculated by two-tailed, paired student’s t-tests. Asterisks indicate p-values as * (p < 0.05), ** (p < 0.01), and *** (p < 0.001).

The currently used monoclonal antibodies (mAb) imdevimab and casirivimab efficiently prevented Delta infection, however, most likely in consequence of amino acid substitutions^5^ failed to neutralize Omicron (**Figure 1D**). In contrast to the currently circulating Delta variant, neutralization efficacy of vaccine-elicited sera against Omicron was severely reduced highlighting T-cell mediated immunity as essential barrier to prevent severe COVID-19. Since Omicron was resistant to casirivimab and imdevimab SARS-CoV-2 genotyping may be needed before initiating mAb treatment. Booster vaccinations, variant-specific vaccines and mAb agents may be required to treat COVID-19 with emerging variants of concern.

## Materials and Methods

### Ethics statement

The study was conducted according to the guidelines of the Declaration of Helsinki, and approved by the Institutional Review Board of the Ethics Committee of the Faculty of Medicine at Goethe University Frankfurt (2021-201, 20-864 and 250719).

### Human sera

Peripheral blood was collected from vaccinated individuals as indicated in **Supplementary Table 4**. All sera were prepared by centrifugation 2000 x g for 10 min, inactivated at 56°C for 30 min, and stored at –20°C until use.

### Virus identification and Sequencing

SARS-CoV-2 isolates were obtained from nasopharyngeal swabs of travel returnees from South Africa as screened by the Public Health Department of the City of Frankfurt am Main, Germany. Swab material was suspended in 1.5 mL phosphate-buffered saline (PBS) and split for RNA-Isolation and viral outgrowth assay. RNA was isolated using the QIAamp Viral RNA Mini Kit (QIAGEN, Hilden, Germany) according to manufacturer’s instructions. RNA was subjected to variant specific RT-qPCR genotyping and Oxford Nanopore sequencing.

### Library preparation, sequencing and bioinformatics analysis

RNA samples extracted from swabs were used for library preparation according to NEBNext ARTIC Standard Protocol (New England Biolabs Ipswich, Massachusetts, USA) (dx.doi.org/10.17504/protocols.io.budxns7n) using the Artic nCoV-2019 V4 primers (IDT, Coralville, Iowa, USA). Libraries were generated using ligation sequencing kit SQK-LSK109, native barcoding expansion kit EXP-NBD104 and FLO-MIN106D R9.4.1 flow cell according to the standard protocol (Oxford Nanopore Technologies, UK) and sequenced on MinION MK1c (Oxford Nanopore Technologies, UK) for 8 h with basecalling and demultiplexing options enabled. The obtained FASTQ files were filtered and analyzed using ARTIC pipeline (https://artic.network/ncov-2019/ncov2019-bioinformatics-sop.html).

See **Supplementary Figure 1** for schematic representation of the SARS-CoV-2 genome indicating spike positions. Sequences are available on GISAID and GenBank under the following accession numbers: SARS-CoV-2 B.1.1.529 FFM-SIM0550/2021 (EPI_ISL_6959871; GenBank ID: OL800702), SARS-CoV-2 B.1.1.529 FFM-ZAF0396/2021 (EPI_ISL_6959868; GenBank ID: OL800703), SARS-CoV-2 B.1.617.2 FFM-IND8424/2021 (GenBank ID: MZ315141).

### Cell culture and Virus Propagation

A549-AT cells^6^ stably expressing ACE2 and TMPRSS2 and Caco2 cells (DSMZ, Braunschweig, Germany, no: ACC 169) were maintained in Minimum Essential Medium (MEM) supplemented with 10% fetal calf serum (FCS), 4 mM L-glutamine, 100 IU/mL of penicillin, and 100 µg/mL of streptomycin at 37°C and 5% CO_2_. All culture reagents were purchased from Sigma (St. Louis, MO, USA).

As described previously SARS-CoV-2 isolates were propagated using Caco2 cells, which were selected for high permissiveness to SARS-CoV-2 infection by serial dilution and passaging as described previously ^7^. Cell-free cell culture supernatant containing infectious virus was harvested after complete cytopathic effect (CPE) and aliquots were stored at −80°C. Titers were determined by the median tissue culture infective dose (TCID_50_) method as described by Spearman^8^ and Kaerber^9^ using Caco2 cells. All cell culture work involving infectious SARS-CoV-2 was performed under biosafety level 3 (BSL-3) conditions. Sample inactivation for further processing was performed with previously evaluated methods^10^.

### Neutralization and antiviral assays

SARS-CoV-2 antibody concentrations were determined using the SARS-CoV-2 IgG II Quant assay and the Alinity I device (Abbott Diagnostics, Wiesbaden, Germany) with an analytical measurement range from 2.98– 5680 binding antibody units per mL (BAU/mL). All sera were initially 1:10 and subsequently serially 1:2 diluted and incubated with 4000 TCID_50_/mL of SARS-CoV-2 Delta or Omicron. Infected cells were monitored for cytopathic effect (CPE) formation 48 h post inoculation. Monoclonal antibody solutions containing imdevimab and casirivimab alone or in combination in equal ratios (1:1) were serially diluted (1:2) and incubated with 4000 TCID_50_/mL of the indicated SARS-CoV-2 variant. After 48 h CPE formation was evaluated microscopically. Evaluation of monoclonal antibodies was quantified using Spark Cyto 400 multimode imaging plate reader (Tecan) as described before^6,11^.

## Data Availability

Sequences are available on GISAID (www.gisaid.org, access date 12/2021), under the following accession numbers. Omicron strains used in this study are as follows: B.1.1.529 (EPI_ISL_6959868; GenBank ID: OL800703), B.1.1.529 (EPI_ISL_6959871; GenBank ID: OL800702). GenBank accession number for the SARS-CoV-2 B.1.617.2 (Delta) isolate IND8424/2021 (GenBank ID: MZ315141).

## Acknowledgements

This study has been performed with the support of the Goethe-Corona-Fund of the Goethe University Frankfurt (MW) and the Federal Ministry of Education and Research (COVIDready; grant 02WRS1621C (MW). We are thankful for the numerous donations to the Goethe-Corona-Fund and the support of our SARS-CoV-2 research. The authors would also like to thank all technical staff involved in data acquisition.

## Supplementary Material

**Supplementary Figure 1.**
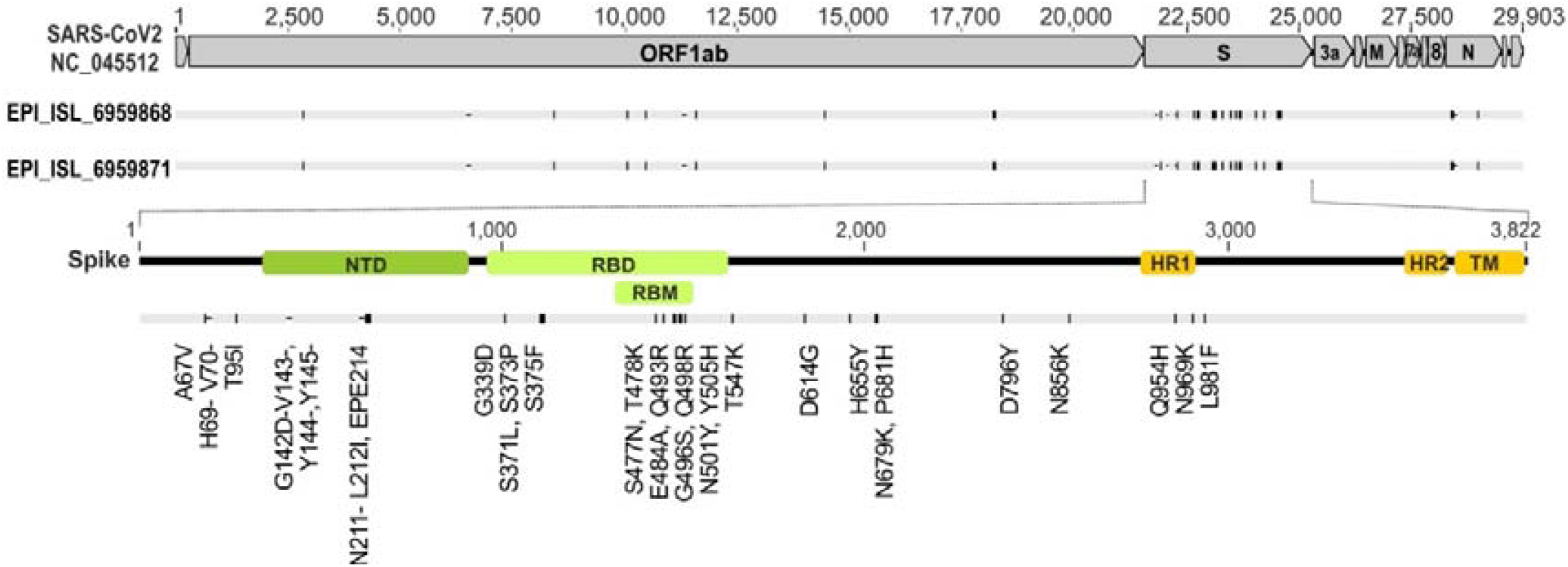
Schematic representation of the SARS-CoV-2 genome indicating spike positions. The numbers denote nucleotide positions based on the reference strain NC_045512. Primary, SARS-CoV-2 Omricon strain FFM-SIM0550/2021 (GISAID: EPI_ISL_6959871; GenBank ID: OL800702) was used in this study. SARS-CoV-2 Omricon strain FFM-ZAF0396/2021 (GISAID: EPI_ISL_6959868; GenBank ID: OL800703) was used for confirmation. NTD RBD and RBM are highlighted by green boxes. HR1, HR2, and TM are indicated by orange boxes. ORF based on reference sequence NC_045512 are shown as grey boxes. Nucleotide substitutions compared to the reference sequence are indicated in the lower section. For sequencing coverage see **Supplementary Table 2**. Dropouts and low coverage regions: Spike: (22796-22983), (23621-23885); E/M gene: (26348-27186). For affected Nanopore Primers see **Supplementary Table 1**.

**Supplementary Table 1.** Dropouts and low coverage Regions: Spike: (22796-22983), (23621-23885) E/M gene: (26348-27186)

| Position in SARS-CoV-2 genome | Affected Nanopore Primer |
| --- | --- |
| 22673 | SARS-CoV-2_76_LEFT |
| 22674 | SARS-CoV-2_76_LEFT |
| 23040 | SARS-CoV-2_76_RIGHT |
| 23048 | SARS-CoV-2_76_RIGHT |
| 23055 | SARS-CoV-2_76_RIGHT |
| 23948 | SARS-CoV-2_79_RIGHT |
| 26270 | SARS-CoV-2_88_LEFT |
| 26577 | SARS-CoV-2_89_LEFT |
| 27259 | SARS-CoV-2_90_RIGHT |

**Supplementary Table 2.**
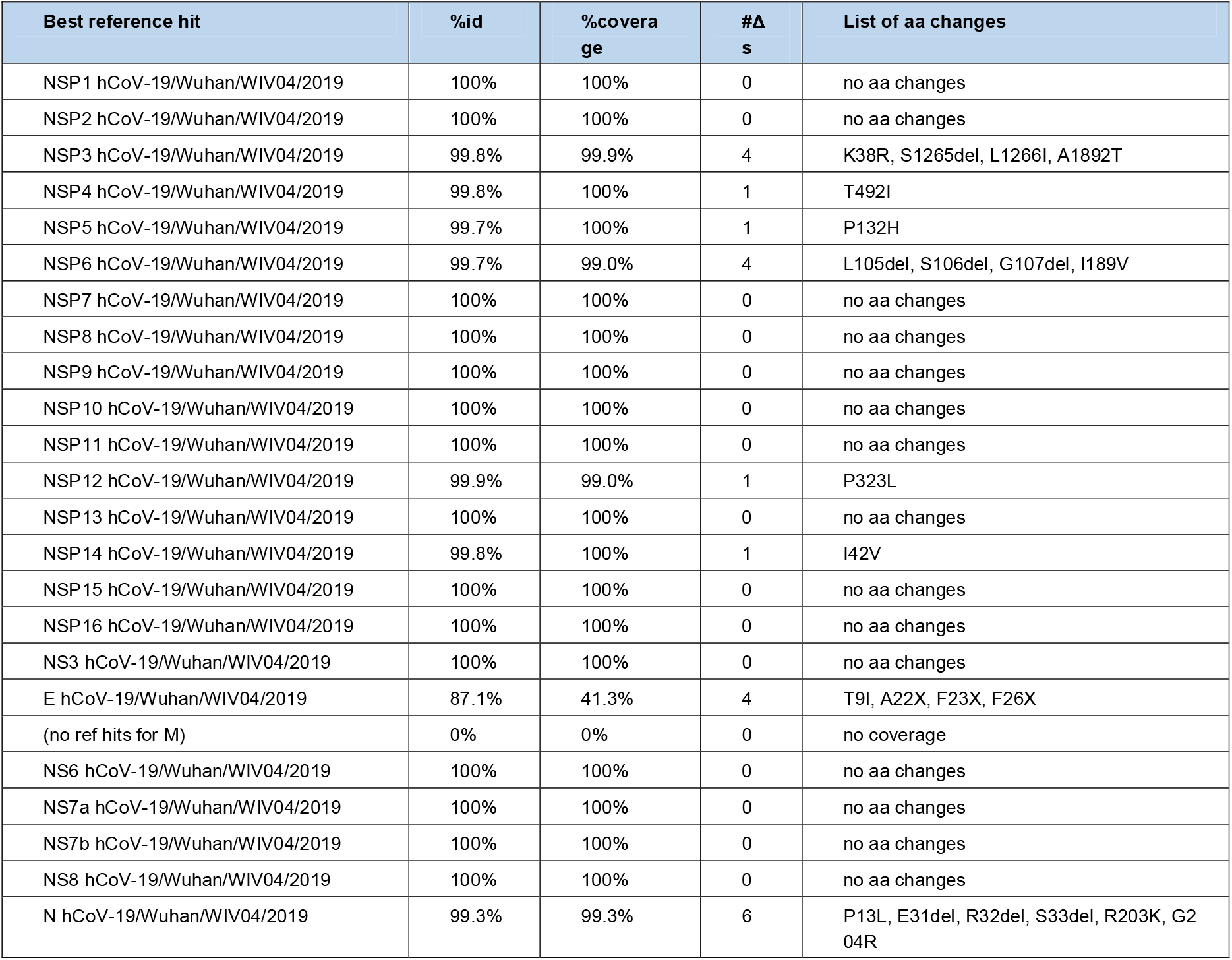
Sequencing coverage, identity and amino acid (aa) changes of SARS-CoV-2 Omicron samples.

| Best reference hit | %id | %coverage | #Δs | List of aa changes |
| --- | --- | --- | --- | --- |
| NSP1 hCoV-19/Wuhan/WIV04/2019 | 100% | 100% | 0 | no aa changes |
| NSP2 hCoV-19/Wuhan/WIV04/2019 | 100% | 100% | 0 | no aa changes |
| NSP3 hCoV-19/Wuhan/WIV04/2019 | 99.8% | 99.9% | 4 | K38R, S1265del, L1266I, A1892T |
| NSP4 hCoV-19/Wuhan/WIV04/2019 | 99.8% | 100% | 1 | T492I |
| NSP5 hCoV-19/Wuhan/WIV04/2019 | 99.7% | 100% | 1 | P132H |
| NSP6 hCoV-19/Wuhan/WIV04/2019 | 99.7% | 99.0% | 4 | L105del, S106del, G107del, I189V |
| NSP7 hCoV-19/Wuhan/WIV04/2019 | 100% | 100% | 0 | no aa changes |
| NSP8 hCoV-19/Wuhan/WIV04/2019 | 100% | 100% | 0 | no aa changes |
| NSP9 hCoV-19/Wuhan/WIV04/2019 | 100% | 100% | 0 | no aa changes |
| NSP10 hCoV-19/Wuhan/WIV04/2019 | 100% | 100% | 0 | no aa changes |
| NSP11 hCoV-19/Wuhan/WIV04/2019 | 100% | 100% | 0 | no aa changes |
| NSP12 hCoV-19/Wuhan/WIV04/2019 | 99.9% | 99.0% | 1 | P323L |
| NSP13 hCoV-19/Wuhan/WIV04/2019 | 100% | 100% | 0 | no aa changes |
| NSP14 hCoV-19/Wuhan/WIV04/2019 | 99.8% | 100% | 1 | I42V |
| NSP15 hCoV-19/Wuhan/WIV04/2019 | 100% | 100% | 0 | no aa changes |
| NSP16 hCoV-19/Wuhan/WIV04/2019 | 100% | 100% | 0 | no aa changes |
| NS3 hCoV-19/Wuhan/WIV04/2019 | 100% | 100% | 0 | no aa changes |
| E hCoV-19/Wuhan/WIV04/2019 | 87.1% | 41.3% | 4 | T9I, A22X, F23X, F26X |
| (no ref hits for M) | 0% | 0% | 0 | no coverage |
| NS6 hCoV-19/Wuhan/WIV04/2019 | 100% | 100% | 0 | no aa changes |
| NS7a hCoV-19/Wuhan/WIV04/2019 | 100% | 100% | 0 | no aa changes |
| NS7b hCoV-19/Wuhan/WIV04/2019 | 100% | 100% | 0 | no aa changes |
| NS8 hCoV-19/Wuhan/WIV04/2019 | 100% | 100% | 0 | no aa changes |
| N hCoV-19/Wuhan/WIV04/2019 | 99.3% | 99.3% | 6 | P13L, E31del, R32del, S33del, R203K, G204R |

**Supplementary Table 3.**
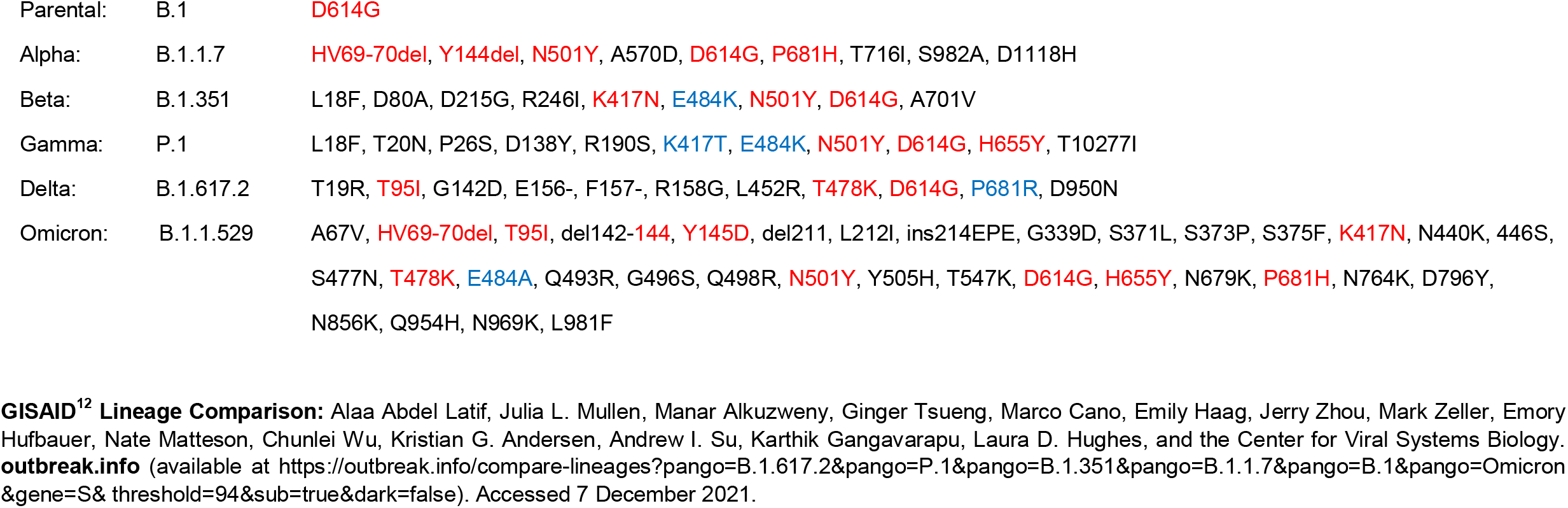
Mutations in the spike proteins of SARS-CoV-2 variants of concern. Compared to the parental SARS-CoV-2 isolate B.1, Omicron gains additional substitutions, insertions and deletions. Amino acid substitutions already found in other variants are highlighted in red. Altered positions but with different distinct substitutions are indicated in blue.

|  |  |  |
| --- | --- | --- |
| Parental: | B.1 | D614G |
| Alpha: | B.1.1.7 | HV69-70del, Y144del, N501Y, A570D, D614G, P681H, T716I, S982A, D1118H |
| Beta: | B.1.351 | L18F, D80A, D215G, R246I, K417N, E484K, N501Y, D614G, A701V |
| Gamma: | P.1 | L18F, T20N, P26S, D138Y, R190S, K417T, E484K, N501Y, D614G, H655Y, T10277I |
| Delta: | B.1.617.2 | T19R, T95I, G142D, E156-, F157-, R158G, L452R, T478K, D614G, P681R, D950N |
| Omicron: | B.1.1.529 | A67V, HV69-70del, T95I, del142-144, Y145D, del211, L212I, ins214EPE, G339D, S371L, S373P, S375F, K417N, N440K, 446S, S477N, T478K, E484A, Q493R, G496S, Q498R, N501Y, Y505H, T547K, D614G, H655Y, N679K, P681H, N764K, D796Y, N856K, Q954H, N969K, L981F |

**GISAID<sup>12</sup> Lineage Comparison:** Alaa Abdel Latif, Julia L. Mullen, Manar Alkuzweny, Ginger Tsueng, Marco Cano, Emily Haag, Jerry Zhou, Mark Zeller, Emory Hufbauer, Nate Matteson, Chunlei Wu, Kristian G. Andersen, Andrew I. Su, Karthik Gangavarapu, Laura D. Hughes, and the Center for Viral Systems Biology. **outbreak.info** (available at https://outbreak.info/compare-lineages?pango=B.1.617.2&pango=P.1&pango=B.1.351&pango=B.1.1.7&pango=B.1&pango=Omicron&gene=S&threshold=94&sub=true&dark=false). Accessed 7 December 2021.

**Supplementary Table 4.** Patient characteristics and overview of sera used in this study.

| Group | Immunization scheme | Group size n | Age Median (range) | Sex (f/m) | BAU/mL Median (IQR) | BAU/mL Mean (range) | months after last vaccination | months after SARS-CoV-2 infection | NT (Delta /Omicron) Median (range) |
| --- | --- | --- | --- | --- | --- | --- | --- | --- | --- |
| 2xBNT<sub>6m</sub> | 2x BNT162b | 15 | 51 (24 - 64) | 11 / 4 | 178 (116.9-290.2) | 289.8 (34.2-1012) | 6 - 7 | 0 | 0 (0-20) / 0(0-0) |
| 2xBNT/<br>BNT<sub>0.5m</sub> | 2x BNT162b +<br>1x BNT162b | 12 | 38 (28 - 59) | 10 / 2 | 2143 (1355-4033) | 2729 (825.3-6250) | 0.5 | 0 | 80 (40-320)/<br>10 (0-20) |
| 2xBNT/<br>BNT<sub>3m</sub> | 2x BNT162b +<br>1x BNT162b | 20 | 43 (26 - 63) | 12 / 8 | 1354 (690.4-2178) | 1838 (167.1-7377) | 3 (+ / - 0.8) | 0 | 20 (0-260)/<br>0 (0-20) |
| 2xMOD<sub>6m</sub> | 2x mRNA-1273 | 14 | 28 (23 - 50) | 7 / 7 | 355 (174-691.1) | 588.8 (90.6-2364) | 6 | 0 | 5 (0-40) /<br>0 (0-0) |
| 2xMOD/<br>BNT<sub>0.5m</sub> | 2x mRNA-1273<br>1x BNT162b | 9 | 29 (23 - 50) | 4 / 5 | 2600 (1879-4089) | 3155 (1422-6540) | 0.5 | 0 | 160 (40-640)/<br>10 (0-40) |
| 1x ChAd /<br>1xBNT<sub>6m</sub> | 1x ChAdOx1<br>1x BNT162b | 19 | 43 (20 - 59) | 14 / 5 | 162.1 (95.5-236.5) | 161.9 (48.6-276.4) | 6 | 0 | 0 (0-10) /<br>0 (0-0) |
| 1x ChAd /<br>2xBNT<sub>0.5m</sub> | 1x ChAdOx1<br>2x BNT162b | 8 | 48 (30 - 59) | 6 / 2 | 1544 (523.9-2931) | 1728 (186.8-3676) | 0.5 | 0 | 60 (0-160) /<br>0 (0-40) |
| 2xBNT /<br>infection | 2x BNT162b<br>+ SARS-CoV-2 | 20 | 87.5 (68 - 93) | 17 / 3 | 1065 (548.7-4407) | 2616 (58.7-11400) | 6 - 7 | 0.7 – 7.6 | 40 (0->1380) /<br>0 (0-160) |

